# Prevalence of Iron Deficiency, Iron Deficiency Anaemia and General Anaemia in Male Gambian Blood Donors Residing in Greater Banjul Region

**DOI:** 10.1101/2021.01.25.21249996

**Authors:** Mustapha Dibbasey, Bolarinde Lawal, Solomon Umukoro, Peter Mitchel

## Abstract

**Objective:** The objective of this study is to determine the prevalence of iron deficiency (ID) and iron deficiency anaemia (IDA) as well as general anaemia in male blood donors and their association with ageing process.

**Methodology and Results:** A total of two hundred and one (201) serum samples were analysed for ferritin in male Gambian blood donors. The ferritin measurement was achieved with COBAS® INTEGRA 400 plus. At the same time, haemoglobin values were retrospectively obtained from the archived haematological full blood count result in the GARIS database. IDA was defined as (Haemoglobin <13.0g/dL+ Ferritin<15ng/ml) whilst ID was defined as (Haemoglobin ≥13.0g/dL+ Ferritin<15ng/ml) and general anaemia was defined as haemoglobin <13.0g/dL in males. The prevalence of anaemia (20%, n=41), ID (22%, n=44) and IDA (10%, n=21), were recorded in male donors. The results show no relationship between ferritin and haemoglobin among the blood donors (collection coefficient (r) = 0.04). Besides, no linear association of having anaemia and ID with ageing was reported among the blood donor population.

**Conclusion and potential application of findings:** ID and IDA as well as general anaemia are highly prevalent among blood donors in the Gambia. Besides, no predisposition to ID and anaemia was observed in term of age, thus all blood donors from 18-60 should be considered for blood donation without any age preference.

## Introduction

Iron is essential in the physiological growth and development of every human being due to its role in oxygen transport, DNA synthesis and electron transport. The most notable and well-established role of iron is manifested in erythropoiesis during haemoglobin synthesis (Abbaspour *et al*. 2014). Haemoglobin is an essential composition of red blood cells (erythrocytes) and composed of haem and globin components which combine to form a complete normal haemoglobin molecule. During erythropoiesis, iron plays a significant role in haemoglobin synthesis as iron is utilised by erythroblasts (the immature erythrocytes) to synthesis haem component. The primary role of haemoglobin is to transport oxygen from the lungs to the tissues (Ganz and Nemeth 2012).

The significant depletion of iron from iron stores coupled with insufficient mobilisation of iron to synthesis haemoglobin, lead to a state of latent ID. Consequently, once iron stores are depleted then iron-deficient erythropoiesis ensues which accompany serious morphological changes on the erythrocytes as they become microcytic and hypochromic. These morphological changes reflect lack of haemoglobin largely due to ID, hence characterised as iron deficiency anaemia (IDA) (Wright *et al*. 2014;Johnson-Wimbley 2011).

ID is the commonest nutritional deficiency in the world. An estimated two billion people worldwide are affected by anaemia mainly due to ID (WHO 2016). The prevalence of IDA is high in low-income countries largely due to insufficient iron intake and loss of blood due to intestinal infection (WHO 2016). In developed countries, certain eating habits (e.g. vegetarianism) and the endemicity of certain pathological conditions (e.g. blood loss or malabsorption) are the most common causes of IDA (de Benoist *et al*. 2005).

Haemoglobin level is used primarily to assess anaemia and is among the worldwide donor selection criteria. WHO defines general anaemia as a haemoglobin level less than 13.0g/dL in males (Johnson-Wimbley 2011). There are several other laboratory tests that are employed to evaluate iron level in humans especially in blood donors. These tests include measurement of ferritin and serum iron (Abbaspour *et al*. 2014).

Measuring ferritin has the potential to accurately assess iron stores in humans as its concentration reduces during the early stage of ID. The normal range for serum ferritin is 30ng/ml to 400ng/ml for males. According to International Nutritional Anaemia Consultative group, individuals above 5 years of age should be regarded as iron deficient when serum ferritin falls below 15ng/dl (WHO 2011). As a result, studies on the prevalence of ID in blood donors have mainly used and recommended ferritin test to establish ID diagnosis in the early stage (Yousefinejad *et al*. 2010; Mittal *et al*. 2006; Erhabor *et al*. 2014). WHO/CDC expert consultation in 2004 recommended haemoglobin and ferritin as the most sensitive markers to detect ID as well as IDA (WHO 2004).

Blood donation is a therapeutic approach used to treat patients diagnosed with severe anaemia (described as Hb≤6.0g/dL). It also has potential benefits for the blood donors: it reduces the risk of acute myocardial infarction in adult men by 88% and lowers the risk of severe cardiovascular events such as strokes by 33% (Salonen *et al*. 1998). The regular blood donation can also improve the efficiency of the donors due to the synthesis of new erythrocytes and reduce the risk of cancer (Zacharski et al. 2008).

However, chronic ID is an established complication that arises mainly due to poorly-regulated blood donation as 225mg of iron is lost for every 450ml volume of blood donated (Jeremiah and Koate 2010). Annually, the maximum number of blood donation that should be achieved by healthy individual blood donors is four times (i.e. every three months) as iron stores can become depleted if the blood is donated more than four times in a year. Studies have shown the high degree of association between the frequency of blood donation (i.e. less than three months intervals) and the risk of developing ID (Cancado *et al*. 2001;Milman and Kirchhoff 1991;Badar *et al*. 2002). As such, studies in different countries have investigated ID in blood donors, an approach to protect regular blood donors and maintain blood supply (Ali *et al*. 2015;Adediran *et al*. 2013;Norashikin *et al*. 2006;Cancado *et al*. 2001;Erhabor *et al*. 2014;Amilo *et al*. 2014;Mittal *et al*. 2006;Mahida *et al*. 2008;Brittenham 2011).

In the Gambia as in most countries the lower age limit is 18 years and upper age limit is around 60 to 70 years. During puberty, both male and female blood donors are at increased risk of ID due to increased iron requirements for growth. Besides, several literatures have reported non-frequent adverse reactions including ID in eligible older donors compared to the younger donors (WHO 2012). In contrast to that, haemoglobin concentration declines with advance in age as ageing process consequently results to progressive and physiological decrement of bone marrow haematopoiesis (Stauder and Thein 2014;Mahlknecht and Kaiser 2010; Alvarez-Uria *et al*. 2014). The impact of growth on the iron stores in the younger population and physiological decrement of bone marrow in adults coupled with the serious lack of regularities on blood donation practice in the Gambia deemed this study worthwhile. In addition, no study is currently done on the prevalence of ID and anaemia in the Gambian blood donors and even in general population (FAO 2010). Hence, this study aims to address prevalence of ID, IDA and general anaemia in the Gambian blood donors and their association with ageing process. This study would help intensify the government approach in protecting potential regular blood donors, which is important in maintaining the reliable supply of blood since the blood bank is constantly experiencing shortage of donors as well as identifying the most appropriate age group for blood donation.

## Materials and Methods

### Study Area

The student study project was conducted in the Medical Research Council (MRC) Clinical laboratories which include Biochemistry and Haematology laboratories department.

### Gambia Adult Reference Interval Study (GARIS)- Study Population

The study was conducted on GARIS samples. The GARIS study is five year project, started in 2013 and expected to end in 2017. GARIS aims to establish haematological and biochemical reference values for Gambian adults. The study targeted male healthy adult blood donors (18 to 60 years) residing in the greater Banjul areas. In the GARIS study, the participants were screened for HIV, VDRL (test for syphilis) and Hepatitis B. The participants with HIV, VDRL or hepatitis B positive were excluded from the study whereas those screened negative for HIV, VDRL and hepatitis B were recruited into the study. The clinical history of the participants was also recorded. However, unfortunately the number of donations performed by the individual blood donors was not captured.

The participants were bled in the three major health centres (Edward Francis Small Teaching Hospital, Brikama Health centre and Serrekunda Health centre). The blood samples were collected by a trained phlebotomist and transported to the clinical laboratories within a 2 hours collection in chilled temperature (2-8°C) to analyse their biochemical and haematological profile.

### Sample Collection

With the permission of the study investigators, the blood samples and haematological data were obtained from the GARIS project. Out of 300 samples, a total of two hundred and one (201) frozen serum samples were obtained for ferritin measurements whilst their haemoglobin and serum iron data were retrieved from GARIS database. The 201 samples with sufficient serum volume were selected (inclusion criteria). All those samples without sufficient serum volume were excluded from the study.

### Sample Analysis

The ferritin measurement were achieved with COBAS® INTEGRA 400 plus (Roche Diagnostics, USA) and the haemoglobin was determined from the Ethylenediaminetetraacetic acid (EDTA) anti-coagulated blood sample using Medonic M-series 3-part haematology analyzer (manufactured by Boule Medical, SWEDEN). Quality controls were run prior to the analysis to ascertain the functionality of both Cobas Integra and Medonic M-series. As part of GARIS, 5ul of serum samples were kept frozen in -20°C for analysis of iron panels which includes ferritin and serum iron. Haemoglobin values were retrospectively obtained from the archived haematological full blood count result in the GARIS database after the ferritin results were already obtained.

### Statistical Analysis

The statistical analysis was performed using SPSS (version 23) and Microsoft Word excel. IDA was defined as (Haemoglobin <13.0g/dL+ Ferritin<15ng/ml) whilst ID was defined as (Haemoglobin ≥13.0g/dL+ Ferritin<15ng/ml) and general anaemia was defined as haemoglobin <13.0g/dL in males. Correlation was performed to investigate whether there is a relationship between age and ferritin concentration as well as between age and haemoglobin level. Then the participants were categorised into three age groups (age group 1: 18-25; age group 2: 26-39; and age group 3: 40-60). Logistic regression was applied to determine an association between ID, and anaemia with the age groups. Odd ratio and 95% confidence interval were determined with logistic regression. Pearson correlation was used to determine correlation between haemoglobin and ferritin level among the donors. A p value of <0.05 denoted a statistically significant.

### Ethical Issues

The GARIS project was approved by the Medical Research Council’s (MRC) Scientific Coordinating Committee and the Gambia Government/MRC Joint Ethics Committee. Written informed consent was obtained from study participants enrolled in the GARIS-study. GARIS participants were assigned unique identification numbers for maintaining anonymity and confidentiality.

## Results

In this study, a total of 201 specimens were considered. The largest percentage of the blood donors were within the age group 26-39 years. The samples were analysed for the presence of anaemia alone using haemoglobin, and for the presence of ID and IDA considering both ferritin and haemoglobin. The prevalence of anaemia (20%, n=41), ID (22%, n=44) and IDA (10%, n=21), were recorded in male donors as shown in table 1. In figure 1, the correlation has shown no association between age and ferritin as well as age and haemoglobin. Among the age groups, the data has shown that individuals within age group 26-39 are about 4 times more likely to have anaemia than individuals within age-groups 18-25 years. However, there is no significant difference in the odds (association) of having anaemia in individuals between 18-25 year olds and 40-60 year olds (in table 2). Besides, no association of having ID among the age groups was found in the study (in table 2). The result of Pearson correlation showed no correlation between ferritin and haemoglobin with a correlation coefficient (r) of 0.04 (Figure 2).

**Table 1:**
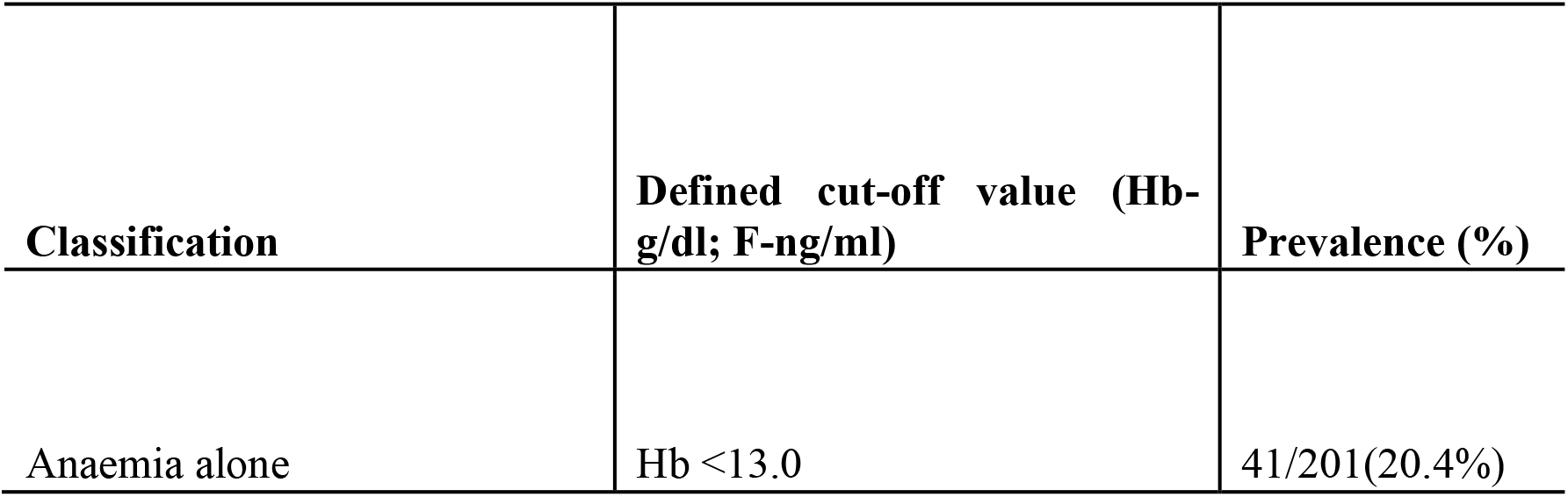
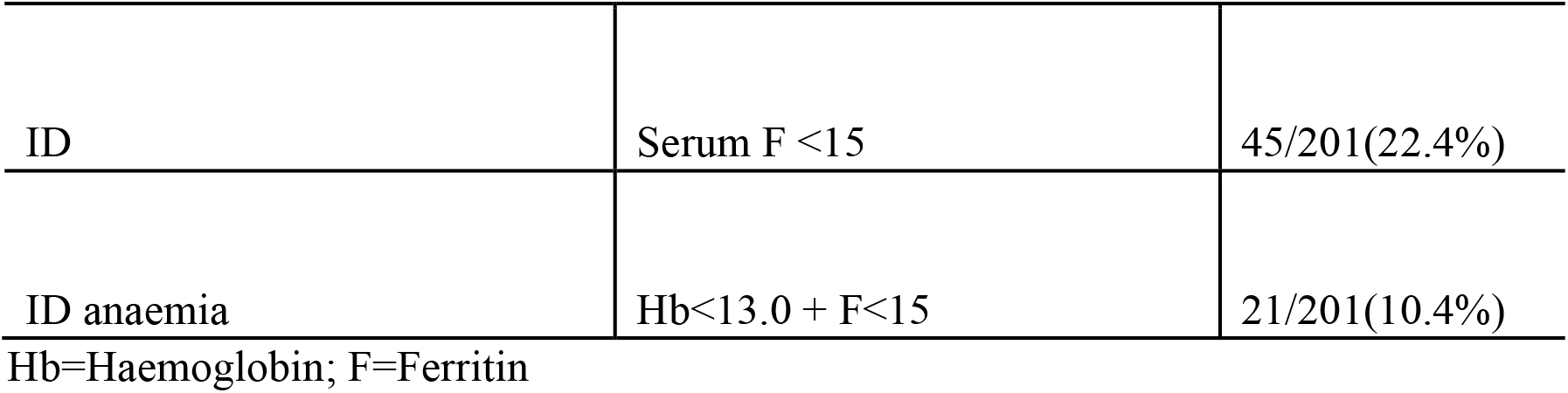
The prevalence of anaemia, ID and IDA among male blood donors recruited in the study

| Classification | Defined cut-off value (Hb-g/dl; F-ng/ml) | Prevalence (%) |
| --- | --- | --- |
| Anaemia alone | Hb <13.0 | 41/201(20.4%) |
| ID | Serum F <15 | 45/201(22.4%) |
| ID anaemia | Hb<13.0 + F<15 | 21/201(10.4%) |

Hb=Haemoglobin; F=Ferritin

**Table 2:** Association of ID and anaemia with age groups

| Age groups | Total (n) | OR | 95% C.I. | P. value |
| --- | --- | --- | --- | --- |
| **Anaemia** |  |  |  |  |
| 18-25 | 21 | 1 |  |  |
| **26-39** | **13** | **4.345** | **1.635-11.548** | **0.03** |
| 40-60 | 7 | .87 | 0.326-2.357 | 0.794 |
| **ID** |  |  |  |  |
| 18-25 | 10 | 1 |  |  |
| 26-39 | 21 | 1.444 | (0.565-3.695) | 0.44 |
| 40-60 | 13 | 1.393 | (0.629-3.085) | 0.41 |

C.I. = confidence interval, OR= odds ratio; 1= Reference; n=total number of anaemia/ID

**Figure 1:**
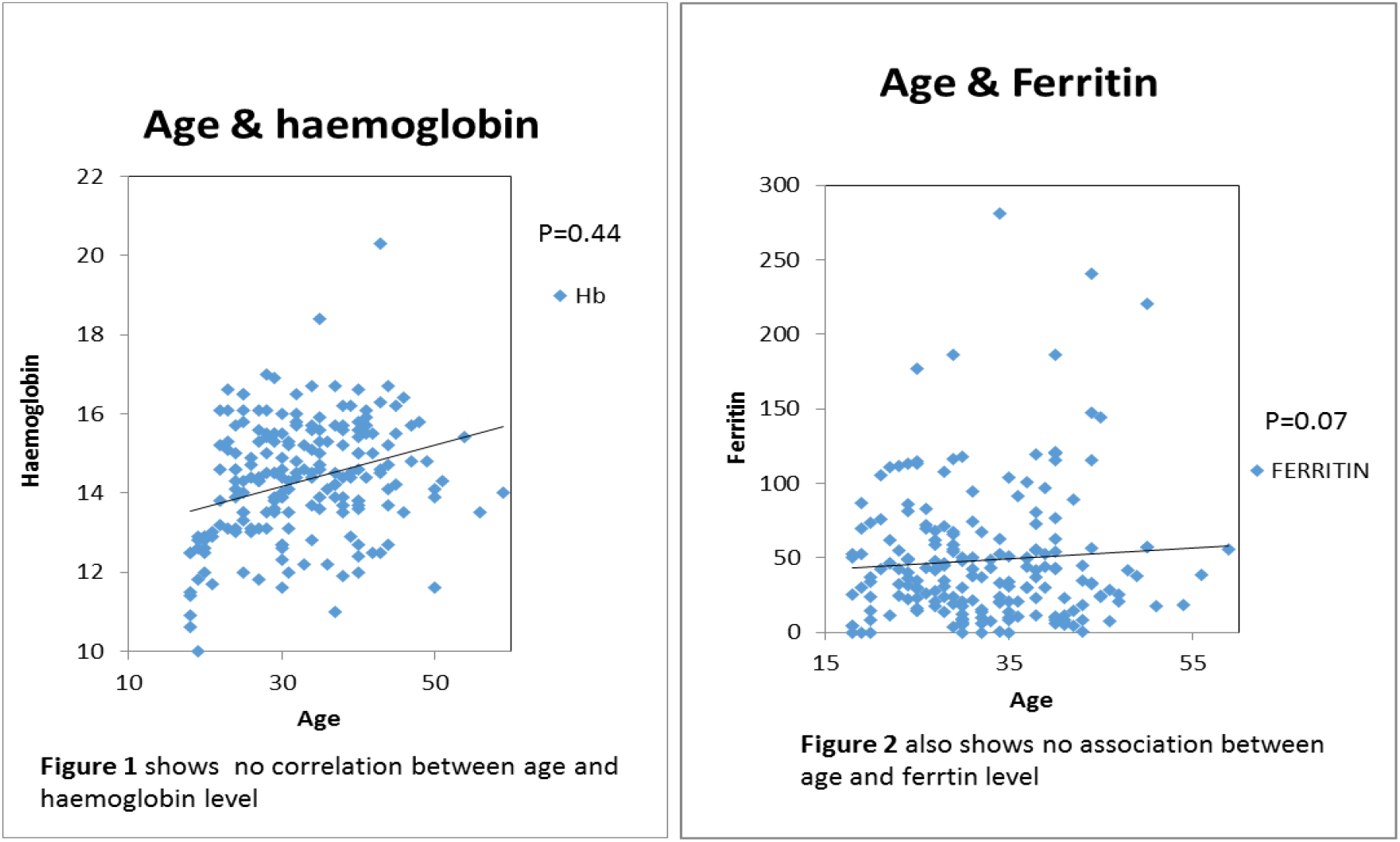
Correlation analysis to investigate whether age can influence the ferritin and haemoglobin levels in the blood donors

**Figure 2:**
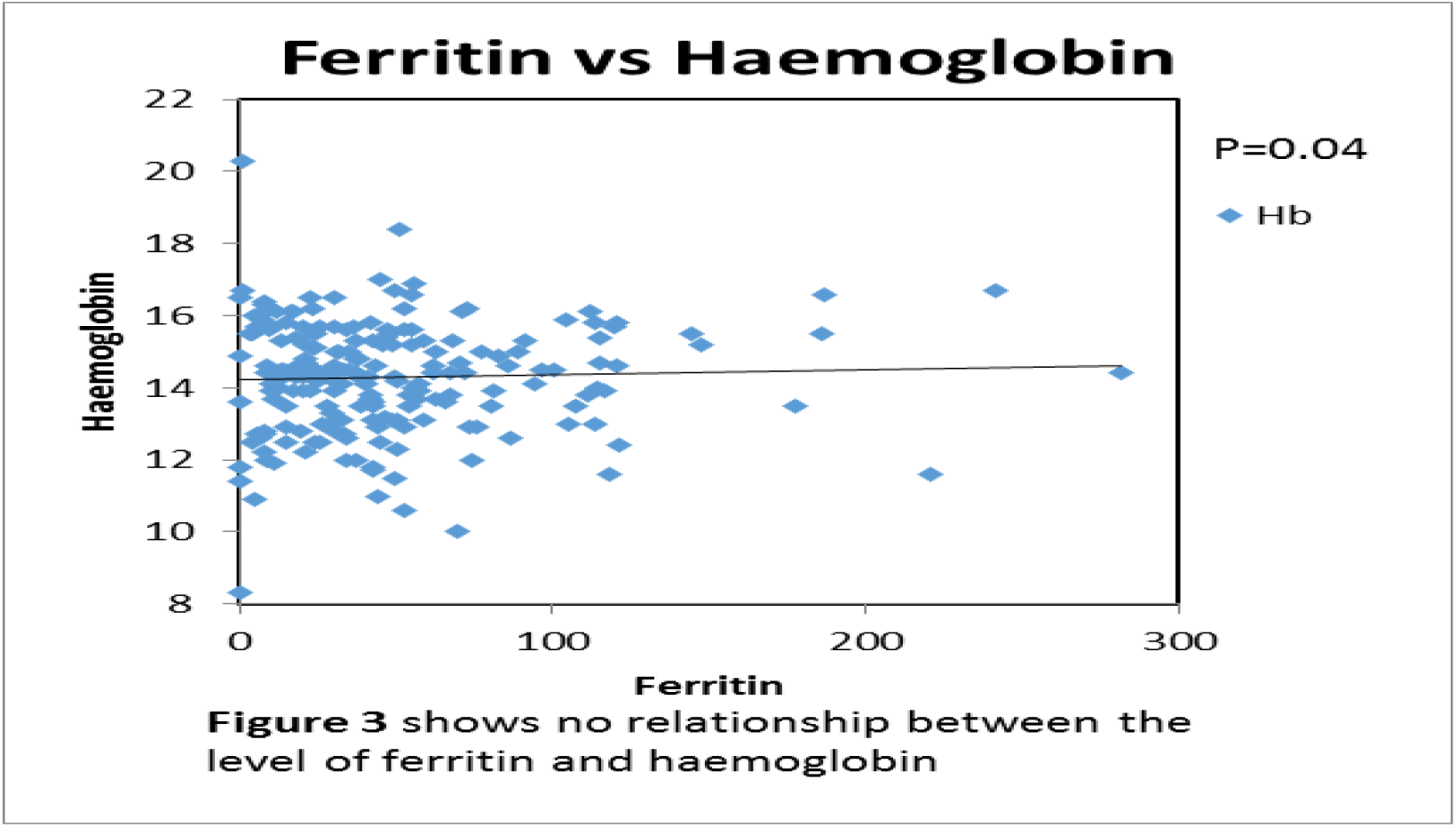
Association (correlation) between Ferritin level and haemoglobin concentration in the healthy male blood donors. The graph has shown that the reduction in iron stores (ferritin) does not reflect a reduction in haemoglobin concentration, hence no association is observed.

## Discussion

The study aims to address the prevalence of ID, IDA and general anaemia among blood donors residing in greater Banjul area and determine their association with ageing process. This is essential to protect the voluntary potential blood donors, maintain the adequate supply of blood in health centres as well as identify the most appropriate age group for blood donation in the Gambia. The study observed a high prevalence of ID of 22.4% among 201 male blood donors. In the Gambia, the nationwide survey on the prevalence of ID centred only on the vulnerable population (in children, pregnant women and breastfeeding women, with prevalence of ID of 23%, 33% and 20% in children, pregnant and breastfeeding women respectively (FAO 2010). As such, no data is available about the ID prevalence in the general population in the Gambia, thus no comparison could be made. However, the finding is consistent with the study of Erhabor *et al*. (2014) conducted in Nigeria which reported 24% prevalence of ID in the blood donors.

Using serum Ferritin and haemoglobin value (Ferritin<15ng/dl+haemoglobin<13.0g/dl), this study reported 10.4% IDA among the male blood donors. Similar trend based on Geneva consultations was noted in this study that the prevalence of ID (22.4%) reported was about twice higher than IDA (WHO 2001;Zimmermann and Hurrell 2007;Alvarez-Uria *et al*. 2014). This finding is in concordance with the findings of studies conducted within Nigeria in West-Africa, 12.0% (Jeremiah and Koate 2010) and 10% (Ali *et al*.2015). However, in our study, the finding of prevalence of ID (22.4%) is higher than the findings of most studies conducted outside West African settings. In Malaysia, Norashikin *et al*. (2006) reported that 11.0% of regular blood donors had ID and in Brazil, study conducted in Sao Paulo also reported 11.0% ID in blood donors (Cancado *et al*. 2001). Also, the finding is ‘by far’ higher than the prevalence of IDA in blood donors reported in some developed countries such as in Iran (2.14%) (Yousefinejad *et al*. 2010) and in Denmark (0.5%) (Milman and Kirchhoff 1991). The high prevalence of ID as well as IDA in blood donors residing in low-income countries including the Gambia is associated with several factors. The most significant contributing factors to high prevalence of ID in developing countries are attributed to low dietary iron intake, parasitic worm infestation, malaria and other infectious diseases such as tuberculosis (de Benoist *et al*. 2005;WHO 2016).

Current practice in the Gambia based on WHO guidelines (WHO 2013) recommends pre-donation haemoglobin of ≥13.0g/dl for males and ≥12.0g/dl for females. Ferritin is highly sensitive to detect iron stores depletion (ID) in humans as its concentration reduces during the early stage of ID. Hence correlation was performed between ferritin and haemoglobin in our study which has shown no correlation with a correlation coefficient=0.07, as demonstrated in **figure: 2**. This is in agreement with the earlier finding that haemoglobin is not a reliable indicator to detect early phase of ID as compared to ferritin (Cancado *et al*. 2001;Jeremiah and Koate 2010). Other factors that can independently cause low haemoglobin concentration with normal or elevated ferritin level includes infections and inflammatory disorders, haemoglobinopathies such as sickle cell disease, vitamin A deficiency and deficiency of folic acid and vitamin B12 (WHO 2007). Similar conclusion was reached in another study conducted in India which studied the correlation between serum ferritin concentration and red cells indices including haemoglobin (Tiwari *et al*. 2012). On the contrary, the finding from our study is not in agreement with the finding of the study of Franchini *et al*. (2007) which reported positive correlation between ferritin and haemoglobin level in 589 male participants. This is likely to be due to the difference in study population as the study of Franchini *et al*. (2007) considered patients with haemochromatosis. Several studies have reported that individuals with primary haemochromatosis have elevated level of erythrocytes parameters including haemoglobin (Jackson *et al*. 2001; Datz *et al*. 1998). Furthermore, Jeremiah and Koate (2010) indicated that some donors may present with latent/early form of ID that may not be detected by measuring haemoglobin level only and may be manifested after a blood donation. In low-income countries, ID causes huge health and economic cost demands that may not be readily achieved (Zimmermann and Hurrell 2007). Thus, early detection of ID is an important tool to reduce the burden of IDA in blood donors which is associated with adverse effects including fatigue, pallor, headache, dizziness, shortness of breath, reduced work performance and intellectual capacity, reduced endurance, impaired immune function, and cognitive changes (Cancado and Laughi 2012). Also, detecting early phase of ID in blood donors is important to the safety and sufficiency of blood supply. ID can be prevented by allowing appropriate donations interval which is sufficient for iron stores restoration, the use of iron supplementation, fortification of processed or staple foods, home food fortification, and increased consumption of food with high iron content and bioavailability such as iron-fortified cereals or bread, brown rice, pulses and beans (Parischa *et al*. 2013;NHS Choices 2016). Hence effective screening of potential voluntary blood donors using ferritin to detect early phase of ID should be efficiently observed at the national blood bank (Yousefinejad *et al*. 2010; Mittal *et al*. 2006; Erhabor *et al*. 2014).

Alongside ID, the prevalence of general anaemia defined by haemoglobin less than 13.0g/dL among the male and 12.0g/dL among the female blood donors was 21.1%. The prevalence cannot be compared to the nationwide survey on the prevalence of anaemia in the Gambia which centred only on the pregnant women (73%), breast-feeding women (56%) and children (76%). Based on WHO criteria, anaemia is a huge public health burden in the Gambia with a prevalence of ≥40% (FAO 2010). The finding is consistent but slightly higher than the findings of Erhabor *et al*. (2014) which reported anaemia prevalence of 16%. The result of anaemia prevalence is also consistent with a study conducted in India which reported that 15.5% of blood donors were deferred due to anaemia (Bahadur *et al*. 2011). The use of different cut-off range to define anaemia with haemoglobin value can impact the outcomes between two different studies. Our result is significantly higher than the result of Jeremiah and Koate (2010) which showed prevalence of anaemia defined by haemoglobin less than 11.0g/dl to be 13.7% (21.1% versus 13.7%). This study as well as other studies have shown that anaemia is a worldwide epidemic problem especially in developing countries and is mainly associated with several factors including nutritional food insecurity complicated by living standard below millennium development gold and high endemicity of infectious diseases such as tuberculosis and malaria (WHO 2016; Stevens et al. 2013; WHO 2015; Gambia Information Site 2016). The high prevalence of anaemia in the Gambia blood donor population is alarming as it could lead to loss of potential, voluntary blood donors. Studies have shown that donor deferred due to low haemoglobin can potentially result to significant loss of donations as donor deferred may failed to return for subsequent donations even though they are eligible candidate (Custer *et al*. 2007). Similar finding was reported in a subsequent study which had shown 50% loss of donors deferred due to low haemoglobin/haematocrit (Custer *et al*. 2010). Consequently, this would lead to insufficient blood supply, and incur high energy and cost in interviewing and recruiting new blood donors. In other to avert this unwanted situation, temporarily deferred donors should be referred for further haematological investigation and treatment. The donors, who have been successfully treated, should be encouraged to return. This study highlight the urgent need to establish donor deferral registry at the National Blood bank to recall the potential blood donors after the deferral period is over (WHO 2012). Better management approach such as iron supplementation, pro-longing donation intervals and changing from whole blood donation to aphaeresis can reduce donor deferral (Pasricha *et al*. 2011; Newman 2008)

### Association of ID and anaemia with ageing

Even though the study presented no linear association of anaemia with age, age group 26-39 was associated with increased odd of having anaemia (odds ratio: 4.35, 95% CI:1.635-11.548, P=0.03) when compared to youngest age group (18-25). This could be explained by uneven distribution of blood donors among age groups; the blood donors in this study are highly concentrated in age-group 26-39 with a population of n=101 as compared to other two age groups (age-group 18-25: n=52 and age-group 40-60: n=48). The finding from the study failed to agree with the finding from the study of Alvarez-Uria *et al*. (2014), which reported that males had a rapid increase in haemoglobin concentrations reaching a plateau of about 14.0g/dL at age 20 years and experienced a progressive decline after age 40 years.

In the case of ID, the result in table 2 showed no association between ID and age substantiated by the correlation analysis between age and ferritin concentration (**shown in the figure 1**), which showed no correlation. However, different outcome was reported in a study conducted in Brazil in which the observed difference only occurred in the blood donors who donated once in the previous year (Moghadam *et al*. 2013). A cross-sectional study showed that age and supplemental iron intake were significantly and positively associated with increased iron stores in women but not in men (Garry *et al*.2000). Hence, this study and the study of Garry *et al*. (2000) have suggested that changes in serum ferritin may not be associated with ageing process in men. The ferritin values for women population were unavailable to confirm positive association between ageing and increased iron stores in women.

The study lacks the ability to address potential confounders which include the information about number of units of blood donated by the blood donors. This information was not captured by the GARIS study too investigate how frequent blood donations impact their iron stores. This could not only be valuable information on the number of blood donation required per individual per year but as a measure to protect voluntary, potential blood donors from a condition of ID. Studies have shown that regular blood donors are highly predisposed to ID than non-regular blood donors (Ali *et al*. 2015; Adediran *et al*. 2013; Norashikin *et al*. 2006; Badar *et al*. 2002; Cancado *et al*. 2001;Amilo *et al*. 2014).

In this study, the information on the eating habit of the blood donors was not available. Since the Gambian diet is predominantly cereal-based, it would be worthwhile to compare the eating habit (vegetarianism) to ID of the blood donors as vegetarianism is among the most causes of ID. This would hence serve as a guideline when offering dietary advice to blood donors (de Benoist *et al*. 2005).

Iron stores are approximately 30% lower in female donors than in male donors who donate once in a year. This is in concordant with findings from studies in which ID is high in female blood donors than in male blood donors (Mahida *et al*. 2008; Cancado *et al*. 2001; Mittal *et al*. 2006; Cancado and Laughi 2012). As a result, the female population was not included in this study due to the fact that only males were recruited into the GARIS, since the study targeted male healthy blood donors.

Gambia like the other countries, the blood donation is predominantly performed by male population. This may negatively impact the applicability of the study outcome in wider population.

In the area of widespread infection, the use of ferritin as single marker to determine ID posts serious challenges. Infection is highly endemic in African settings including the Gambia. However, no data such as white blood cell count is available to support this finding in blood donors in this study (Barreto *et al*. 2006). Hence, this might impact the overall prevalence of ID in blood donors reported in this study. The future large-scale studies on the prevalence of ID in blood donors should consider acute phase proteins such as C-reactive protein and acid-glycoprotein alongside ferritin to aid in the accurate interpretation of ferritin values since ferritin level is elevated in inflammation/infection despite low iron stores (WHO 2011). Since is an acute phase protein, ferritin level can be elevated concurrently in patients with either ID, and chronic inflammation, malignancy or liver disease (Mukhopadhyay and Mohanaruban 2002).

ID and iron-deficiency anaemia as well as anaemia are highly prevalent among blood donors in the Gambia. Besides, no predisposition to ID and anaemia was observed in terms of age, thus all blood donors from 18 to 60 should be considered for blood donation without any age preference. However, this might not be conclusive as the number donations, which is not available, might have impacted the outcome of this study. Haemoglobin value is not a reliable tool to predict ID in blood donors. Hence the study highlights the need to review the screening tests for the selection of newly recruited blood donors and include serum ferritin measurement for the routine assessment of regular blood donors. The study highlights the need for nationwide coverage to include blood donors across the country with comprehensive demographic details and clinical history of infection and number of donations made in a year. Female population should also be considered in this coverage if possible.

## Supporting information

Ethical Approval form

## Data Availability

This is a simple prevalence study to provide an insight into the burden of iron deficiency in Gambia blood donors. This a sub-study for GARIS that gained approval from the Ethics Committee.

## Acknowledgments

I would like to acknowledge the Clinical Laboratories Manager (Bolarinde Lawal) for granting me the access to GARIS samples and database as well as provision of ferritin reagents. I would like to acknowledge both the e-tutor (Susan Clarke) and workplace supervisor (Gibril Bah) as well as Dr Brenda Kwambana for their intellectual contributions and feedbacks.

## Abbreviations list

(ID): Iron deficiency
(IDA): Iron deficiency anaemia
(GARIS): Gambia Adult Reference Interval Study
(HIV): Human Immunodeficiency Virus
(VDRL): Veneral disease Research Laboratory
(MRC): Medical Research Council
(SPSS): Statistical Package for Social Science
(WHO): World Health Organisation
(ANOVA): Analysis of Variance
(EDTA): Ethylenediaminetetraacetic acid

## Notes

### Competing Interest Statement

The authors have declared no competing interest.

### Clinical Trial

The study was simple prevalence study under the main study known as GARIS study that aimed to establish the reference range for Gambia adults.

### Funding Statement

No external funding received for this studies

### Author Declarations

The Gambia Government/MRC Joint Ethics Committee provided approval to GARIS study. The study is a sub study of GARIS project that established reference intervals of haematological and clinical chemistry profiles in apparently healthy Gambian Adults.

