## Supplementary material for "Prevalence of Iron Deficiency, Iron Deficiency Anaemia and General Anaemia in Male Gambian Blood Donors Residing in Greater Banjul Region": Ethical Approval form

The Gambia Government/MRC Joint
**ETHICS COMMITTEE**

C/o MRC Unit: The Gambia, Fajara
P. O. Box 273, Banjul
The Gambia, West Africa

3 February 2014

Mr Ameh James
Clinical Laboratories Manager
Disease Control and Elimination Theme
MRC Unit, The Gambia
Fajara

Dear Mr James

**SCC 1360v2, Haematological and clinical chemistry profile in apparently healthy Gambian Adults: in search of reference intervals**

Thank you for submitting the revised participant information sheet as requested by the Gambia Government/MRC Joint Ethics Committee at its meeting held on 29 November 2013.

I am happy to record our Committee's full approval for the commencement of this study.

With best wishes

Yours sincerely

Mr Malamin Sonko
Chairman, Gambia Government/MRC Joint Ethics Committee

**Additional documents submitted for review:-**

- Participant Information Sheet, Version 2.0 – 25 January 2014
- Consent Form, Version 2.0 – 25 January 2014
- Questionnaire
- CV-Ameh James

**The Gambia Government/MRC Joint Ethics Committee:**

*Mr Malamin Sonko, Chairman*
*Professor Ousman Nyan, Scientific Advisor*
*Ms Naffie Jobe, Secretary*
*Mrs Tulai Jawara-Ceesay*
*Dr Ahmadou Lamin Samateh*
*Dr Roddie Cole*
*Professor Tumani Corrah*
*Dr Stephen Howie*
*Dr Kalifa Bojang*
*Dr Ramatoulie Njie*
*Dr Adama Demba*
*Dr Siga Fatima Jagne*
